# The impact of extreme temperatures on birth outcomes in the Netherlands: a nationwide population-based study

**DOI:** 10.1101/2023.08.15.23294111

**Authors:** Lizbeth Burgos Ochoa, Pilar Garcia-Gomez, Eric AP Steegers, Tom Van Ourti, Loes CM Bertens, Jasper V Been

## Abstract

**Background:** Climate projections predict an increased frequency and intensity of extreme weather events, such as extreme temperatures, prompting concerns about their impact on early-life health and health disparities. This study aimed to investigate the causal impact of in-utero exposure to extreme temperatures on birth outcomes and effect heterogeneity across levels of socioeconomic status (SES).

**Methods:** We obtained data on singleton births that occurred between 2003 and 2017 from the Dutch perinatal registry (N=2 472 352). Exposure was calculated as the number of days during the gestational period in which the mean temperature fell into each of mutually exclusive bins, with the 8–12 °C bin used as reference. To identify a causal effect, we exploited the unpredictability of daily temperature fluctuations while accounting for seasonal and underlying trends. Effect heterogeneity was assessed across levels of household income, neighbourhood SES, and maternal education.

**Results:** In-utero exposure to an additional hot day (mean temperature > 20 °C), relative to the reference range, led to increased odds of low birth weight (LBW) (OR[95%CI]= 1.007 [1.005, 1.009]), small for gestational age (SGA) (OR[95%CI]= 1.004 [1.003, 1.005]), and preterm birth (PTB) (OR[95%CI]= 1.006 [1.005, 1.007]). Exposure during the second trimester to an additional cold day (< -4 °C) led to increased odds of LBW and PTB. The observed effects were the most detrimental for births in low-SES households.

**Conclusions:** In-utero exposure to extreme temperature has a detrimental impact on birth outcomes in the Netherlands. Projected increases in extreme temperatures may further exacerbate health disparities in early life.

## BACKGROUND

The latest assessment by the United Nations’ Intergovernmental Panel for Climate Change highlights that climate change has increased the intensity and frequency of extreme weather events such as extreme temperatures.^1^ This increase is likely to be especially burdensome for vulnerable populations with limited capacity to respond or adapt to extreme weather events.^2^ Concerns about health effects drive climate policy focus, prompting global calls for more research to aid adaptation strategies for at-risk groups.^3–5^

Birth outcomes, such as low birthweight (LBW), small-for-gestational-age (SGA), and preterm birth (PTB) have been recognized as key influential factors in a child’s life-course development and health.^6^ There is a substantial body of literature showing that adverse environmental exposures during the gestational period affect health outcomes at birth;^6^ but also have long lasting effects on the health, educational and economic outcomes of the affected children.^6,7^ From a biological point of view, foetuses are likely affected by extreme temperatures because of the physiological changes that alter mothers’ capacity to regulate body temperature.^4,8,9^ Furthermore, animal models support the biological plausibility of a detrimental relationship between extreme temperature and health at birth.^10,11^

The available literature mostly consists of association studies on temperature and birth outcomes. Although inconsistently, these studies provide evidence that exposure to extreme temperatures is associated with higher risk of adverse birth outcomes.^5,12^ However, many lack accounting for critical confounders, hindering causal interpretation. Methodological disparities and exposure definitions further complicate synthesis and comparison.^12–14^ For example, a major criticism of earlier studies is the reliance on the concept of ‘heat waves’ which lacks a universal definition.^13,15,16^ Recently, a small number of studies, mostly from the field of economics, have been able to establish a causal relationship between extreme temperatures and health at birth.^17–22^ These studies used a “binned” approach to modelling the temperature-response function which allows for nonlinear effects of temperature on health outcomes, facilitates the control for sources of confounding, and is suitable for different types of outcomes.^23^ This approach has a clear definition of exposure, i.e., the number of days in the gestational period with temperatures falling within prespecified degree ranges (bins).^23^ Additionally, it enables examination of effects from both temperature extremes, particularly relevant as there have been mixed findings on health impact of hot and cold temperatures.^24^ Findings mainly support extreme high temperatures’ impact on LBW, but other outcomes are understudied.^17,18,21,22^ LBW can arise from preterm delivery, intrauterine growth restriction (SGA), or a combination.^25^ As extreme temperatures affect SGA and PTB through distinct mechanisms,^10,26^ causal evidence is crucial. Yet, such evidence for SGA and PTB remains insufficient.^19^

Given the projected rise in extreme weather events, comprehending the impact of extreme temperatures on birth outcomes and its variation by socioeconomic status (SES) is crucial.^27^ Yet, despite some exploration, existing research lacks a consensus on the socioeconomic heterogeneity of extreme temperature effects on birth outcomes.^13^ SES may influence these effects through factors like housing conditions, workplace disparities, risk behaviour awareness, and access to mitigation measures.^13^

The objective of this study is to investigate the causal effect of extreme temperatures on key adverse birth outcomes in the Netherlands. Our work expands previous literature by assessing whether any observed impact on LBW could be due to preterm delivery, intrauterine growth restriction, or both. Furthermore, we examine the role of socioeconomic status (SES) as moderator for the effect of extreme temperatures on birth outcomes. Results from this study may inform the development and optimization of existing adaptation strategies and management of pregnant women during and after extreme temperature periods.

## METHODS

This is a national retrospective study based on individual-level birth records linked to routinely collected climatological data and population register data. The study comprises singleton births at gestational ages between 24+0 and 41+6 weeks that occurred in the Netherlands between 1 January 2003 and 31 December 2017.

### Data sources

Birth records were obtained from the Netherlands Perinatal Registry (Perined), which includes more than 97% of all deliveries in the Netherlands. Perined provides individual-level information on pregnancy and birth outcomes, along with maternal characteristics, and the four-digit postcode of the mother’s place of residence at delivery. Additionally, linkage of Perined to Statistics Netherlands (CBS) records was performed to retrieve maternal and household sociodemographic information. However, 3% of the births recorded in Perined could not be linked by CBS (including stillbirths). Detailed information about the linkage procedures can be found at the CBS website.^28^

Data on meteorological conditions was obtained from the Royal Netherlands Meteorological Institute (KNMI).^29^ Meteorological data is collected by KNMI using several monitors placed across the entire country. The KNMI data informs on daily mean (along maximum and minimum) ambient temperature in °C, total precipitation (in millimetres), wind speed (in meters per second), and sunshine duration (in hours). We matched each birth record to daily weather records during the full gestational period from the nearest monitor to the place of mother’s residence (postcode). The average matching distance is 15 km, which is smaller than the one observed in previous studies.^18,19^

### Variables

The study outcomes are the following: 1) low birth weight (LBW), i.e., birth weight below 2,500 grams, 2) Small-for-gestational-age (SGA), i.e., birth weight below the 10th centile adjusted for gestational age and sex, according to national reference curves,^30^ and 3) preterm birth (PTB), i.e., birth occurring before 37+0 weeks.

To facilitate comparison with previous studies, the exposure was set as the number of days during the gestational period in which the daily mean temperature falls into each of mutually exclusive temperature bins i.e., < -4 °C, -4 – 0 °C, 0 – 4 °C, 4 – 8 °C, 8 – 12 °C, 12 – 16 °C, 16 – 20 °C, and > 20 °C.^18,19,31^ Higher temperature bins were considered, i.e., up to > 28 °C. However, the exposure during the gestational period to days with a mean temperature >28 °C was on average only 1.4 days (in comparison to the 19.0 days observed in Chen et al.^19^), often leading to very wide confidence intervals for higher bins. The gestational period was determined using the birth date and gestational age, which was used to calculate the date of conception.^32^ Gestational age, obtained from the Perined dataset, is estimated by the healthcare provider using information on the last menstrual cycle and foetal scans to ensure accuracy.^33^

The linkage of Perined with CBS microdata allowed access to a set of sociodemographic variables. Information on equivalized household disposable income during the year of birth (corrected for size and composition of the household)^34^ was categorized into low, medium and high where the low and high categories correspond to the lowest and highest quintiles, respectively. Mother’s highest educational level is classified by CBS as low, medium, high, or unknown.^35^ Moreover, we obtained maternal migration background as defined by CBS based on country of birth, i.e., Dutch, Turkish, Moroccan, Surinamese, Antillean, others western, and others non-western.^36^ We used SCP Status Scores to assess neighbourhood socioeconomic status.^37^ These scores combine yearly data on income, employment, and education for four-digit postcodes. SES categories were established using quintiles from the Status Scores: lowest and highest for low and high SES, and the middle for medium SES.

### Empirical strategy and challenges to causal effect identification

To estimate the effect of ambient temperatures on birth outcomes we used logistic regression models. We used a “binned” approach to model the temperature-response function,^23^ where the bin 8–12 °C (which includes the yearly average temperature in the Netherlands), was excluded and used as reference category in all models.

Studying temperature’s causal impact on birth outcomes is complex due to non-random exposure and correlations with outcomes can emerge without causation. Some studies observe health differences in children born in different months due to parental characteristics and seasonal factors (e.g., influenza virus) influencing conception.^38,39^ Moreover, regional geographic characteristics may also be correlated with both weather and outcomes.^40^ Accordingly, our models control for differences in outcomes due to seasonality, regional variation, and time trends by including province × (conception) month fixed effects, a province × linear year-time-trend, and year fixed effects. Moreover, a broad set of climatological control variables was included in the models, i.e., the average precipitation, sunshine duration, and wind speed. The models were not adjusted for mediators such as ambient air pollution since we are interested in the total effect of ambient temperature on birth outcomes.^41,42^

In the literature, another concern is families sorting into warmer or cooler regions based on sociodemographic characteristics and preferences. However, the Netherlands has a mild maritime climate, historically marked by mild summers and rare excessive heat. Uniform temperature due to the flat landscape means provinces differ by just around 1°C.^43^ With these slight climate differences, non-random sorting of pregnant women is less likely. Temperature’s unpredictability suggests exposure’s independence from maternal traits after considering seasonality and trends. We test this assumption in sensitivity analyses.

Prior research has noted a mechanical correlation between gestation length and likelihood of exposure, leading to spurious associations as children with longer gestations have a longer time in which they could be exposed.^32^ To overcome this issue, we followed the approach by Currie and colleagues,^32^ where the exposure is constructed using a hypothetical gestational period (counting 280 days forward from the day of conception) instead of the actual length of gestation. This approach, is now standard in the literature looking at the impact of environmental in-utero exposures.^44^ Survivor bias was addressed by including covariates linked to outcomes and early foetal loss^45^: maternal age in categories (≤19, 20-34, ≥35 years), parity (nulliparous vs multiparous), foetal sex, (equivalized) disposable household income, mother’s educational level, and maternal migration background.

To explore SES moderating extreme temperature’s impact on birth outcomes, we included interaction terms between temperature bins and SES indicators. The primary indicator was equivalized disposable household income (low, medium, high). Additional analyses were conducted for mother’s education and neighbourhood SES.

To assess susceptibility windows to extreme temperatures, we examined trimester-specific exposures. Using the calculated date of conception, weeks 1–13 after conception date were assigned to trimester 1, weeks 14–26 to trimester 2, and week 27 and above to the third trimester.^19,46^ Given that some births occur before the third trimester it is possible that our results for this trimester would be biased downwards, particularly for PTB (8% of PTB deliveries occur before the third trimester). Our approach treats these cases as if they were still at risk; however, under the rare disease assumption (prevalence <10%), it has been shown that any bias due to these sorts of strategies is minimal and generally negligible.^47^

To assess the validity of our results, an extensive set of sensitivity analyses was conducted. First, we conducted analyses with a negative control exposure (placebo test) to detect bias linked to residual unobserved confounding due to non-random sorting,^48^ i.e., we used temperature exposures corresponding to 9 months after the birth instead of the actual exposure.^19^ To address measurement error, models were additionally adjusted for distance to the monitor location; if distance leads to measurement error, this strategy may help reducing the bias. We assessed the heterogeneity of the results according to foetal sex by including interaction terms between exposure and sex. Finally, in a similar fashion to previous studies, we also conducted analyses using maximum and minimum temperature for the exposure bins. All analyses were conducted in R version 4.0.6. ^49^

## RESULTS

Between 2003 and 2017, 2 629 207 births were registered in the Netherlands. After removing multiple births, births with gestational age below 24+0 weeks or above 41+6 weeks, and births with missing data on covariates, there were 2 472 352 births available for the main analysis. Summary characteristics of the population are shown in table 1. The prevalence of LBW, SGA and PTB were 4.7%, 11.4%, and 5.8%, respectively. Also, Table 1 shows the distribution of average number of days during the gestational period falling into each of the temperature bins. On average, pregnant women during the study period were exposed to 12.4 days with a mean temperature falling > 20°C and 2.5 days corresponding to the range < -4 °C.

Figure 1 shows the estimates for the effect of in-utero temperature exposure on birth outcomes (numerical results available in supplementary file 1). In-utero exposure to an additional hot day, i.e., with mean temperature > 20 °C, relative to a day within the 8 – 12 °C range, was related to increased odds of LBW (OR[95%CI]= 1.007 [1.005, 1.009]), SGA (OR[95%CI]= 1.004 [1.003, 1.005]) and PTB (OR[95%CI]= 1.006 [1.005, 1.007]). There was also a detrimental effect of exposure to an additional day in the 16 – 20°C range that was smaller in magnitude (see Figure 1). The point estimates of exposure to an additional cold day throughout the full gestational period showed a detrimental effect for LBW and PTB, however the confidence intervals covered the null. Concerning the timing of the exposure, we observed that in all trimesters exposure to an additional day > 20 °C (relative to the reference) had a detrimental impact for SGA while for LBW and PTB an effect was only observed in the second and third trimesters (supplementary file 2). Regarding cold temperatures, we found that exposure during the second trimester to an additional day with mean temperature < - 4 °C (relative to the reference) had a negative impact on LBW and PTB, but not on SGA.

**Figure 1.**
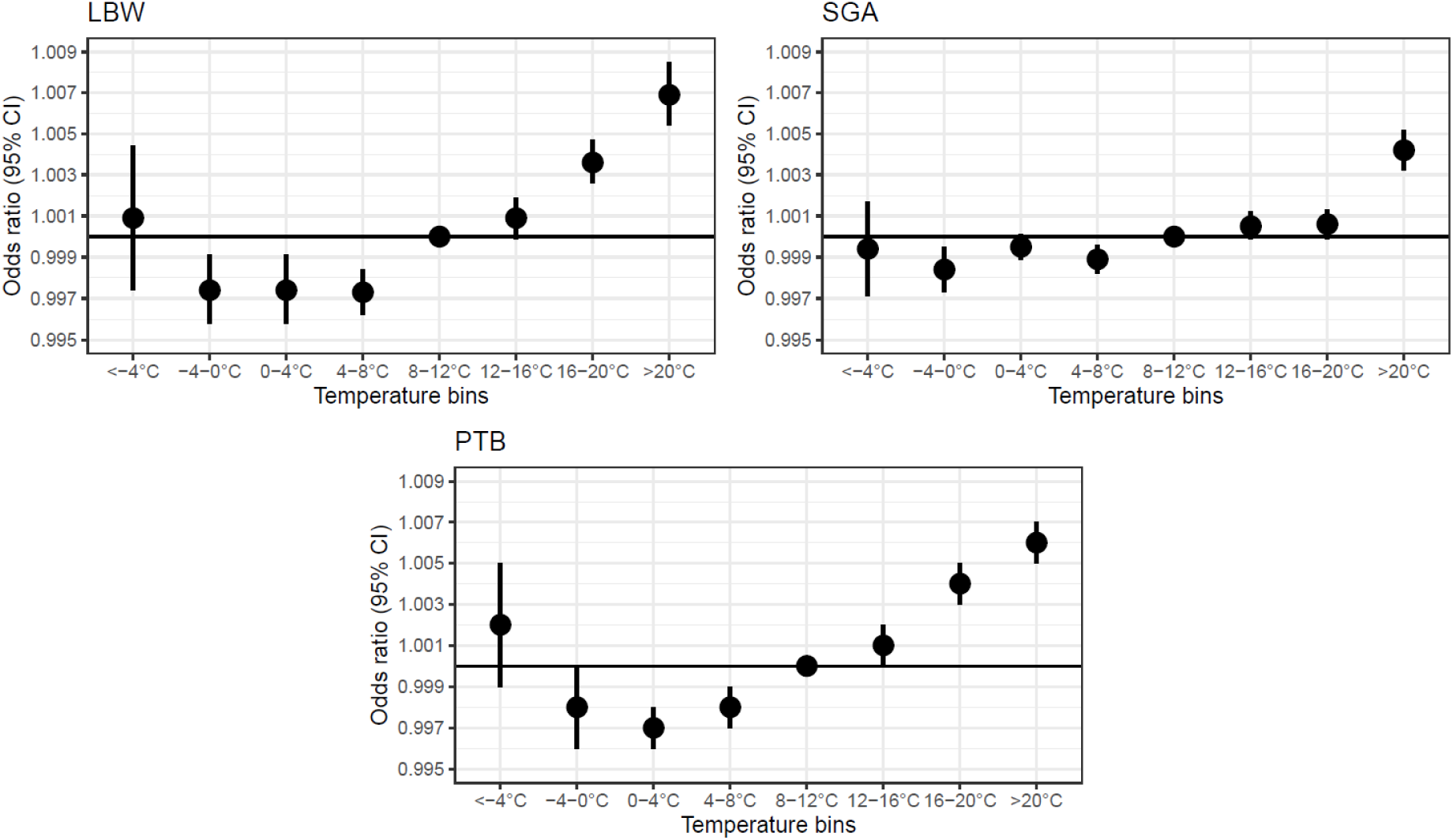
The effect of in-utero exposure to one additional day falling in certain temperature bin on birth outcomes (relative to a day with a mean temperature of 8 – 12 °C). **Footnote:** All models include province × week-of-the-year fixed effects, province × year-time-trend, and year fixed effects. Environmental controls include mean precipitation, wind speed, sunshine duration, and relative humidity. Other covariates included were maternal age in categories, parity, fetal sex, household income, mother’s migration background and education.

**Figure 2.**
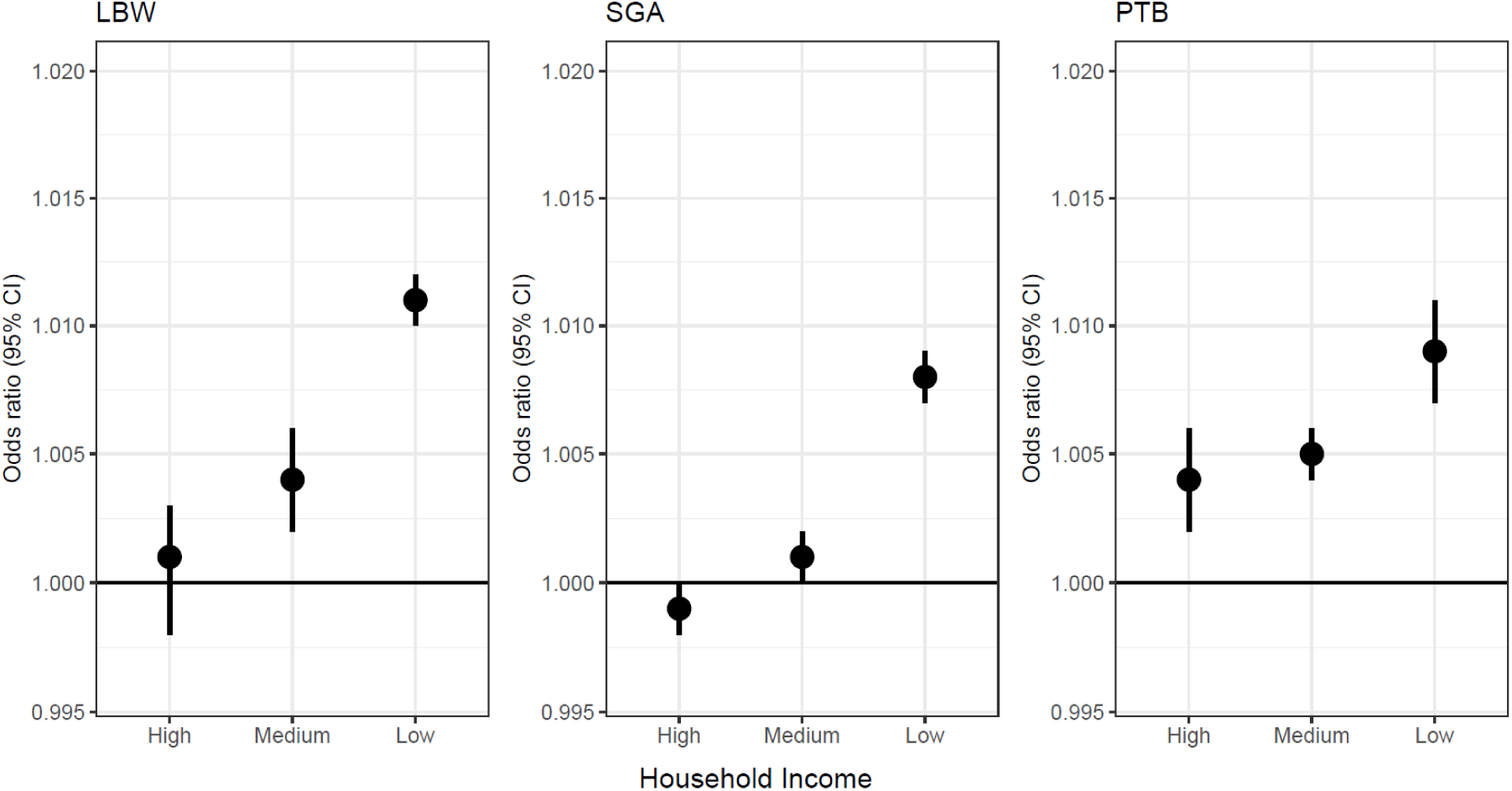
The effect of in-utero exposure to one additional hot day (>20 °C) (relative to a day with a mean temperature of 8 – 12 °C) by household income. **Footnote:** All models include province × week-of-the-year fixed effects, province × year-time-trend, and year fixed effects. Environmental controls include mean precipitation, wind speed, sunshine duration, and relative humidity. Other covariates included were maternal age in categories, parity, fetal sex, household income, mother’s migration background and education.

**Table 1.**
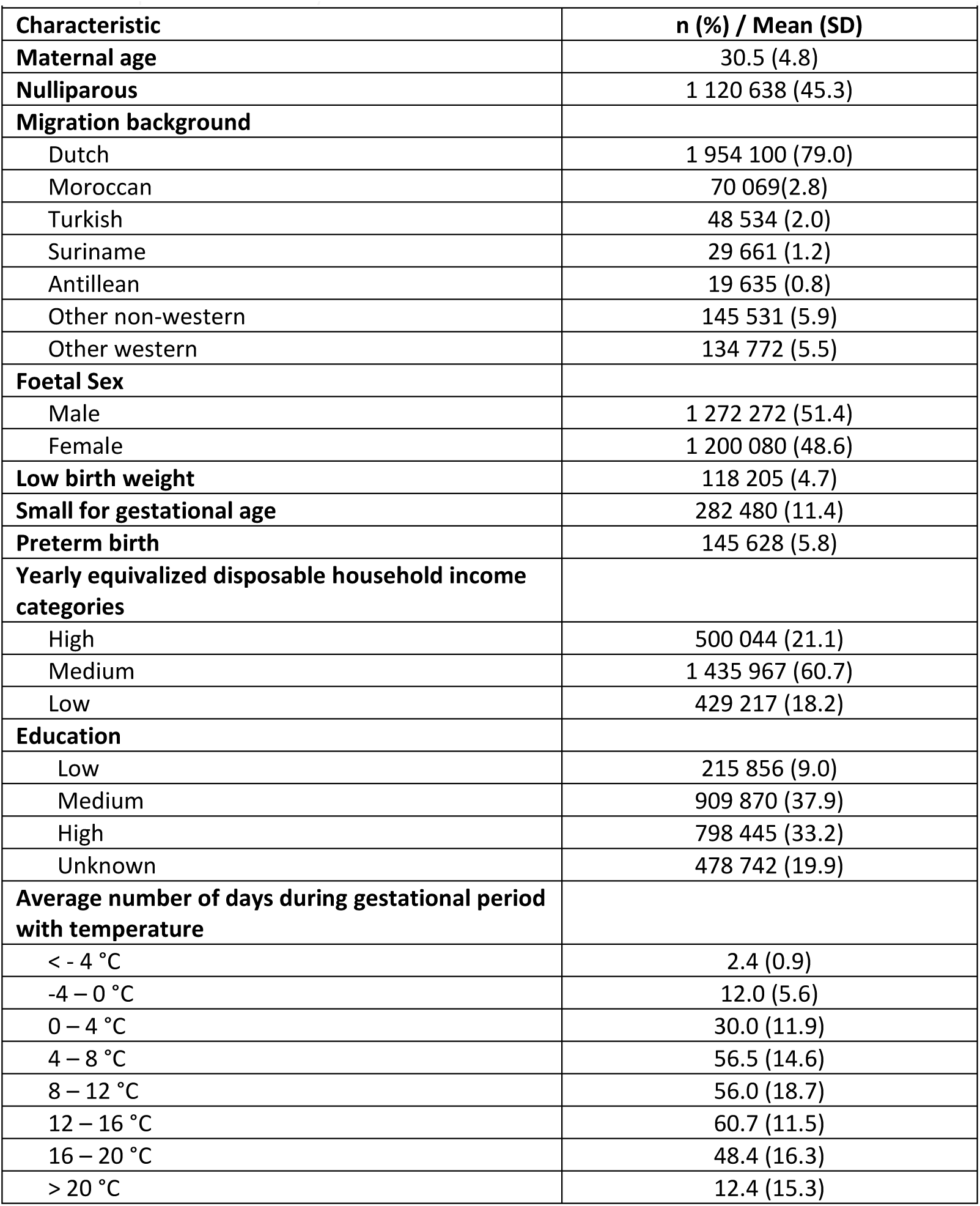
Population summary characteristics.

We observed that the detrimental impact of in-utero exposure to extreme temperatures was more pronounced for births in low-income households. Some of the largest differences were observed in SGA, e.g., the effect of an additional day > 20 °C for low-income households corresponded to OR(95%CI)= 1.013 (1.012, 1.014) while for high income households this was OR(95%CI)= 0.998 (0.997, 0.999). Similarly, for cold temperatures, we found that the effect of an additional day < -4 °C had a detrimental effect on births from low-income households (OR[95%CI]= 1.016 [1.012, 1.019], however, this was not the case for high income households (OR[95%CI]= 0.989 [0.986, 0.992]). Similar patterns were found in the analyses including interaction between exposure and neighbourhood SES (supplementary file 3). However, less heterogeneity was found for maternal education.

Patterns from the main analysis held in sensitivity tests adjusting for distance to monitor location. Consistent patterns emerged using daily maximum and minimum temperature, with larger effects seen in maximum temperature. The absence of observed patterns in the placebo test suggests self-selection based on unobserved traits is unlikely. Additionally, extreme high temperatures had a more negative impact on males than females, especially for PTB. See supplementary file 4 for detailed results.

## DISCUSSION

In this nationwide study in the Netherlands, we found evidence of a negative impact of in-utero exposure to extreme ambient temperatures on key birth outcomes. We consistently observed that an additional day with maximum temperature > 20 °C increased the odds of LBW, SGA, PTB. We also found a detrimental effect of exposure to cold temperatures (< -4° C) during the second trimester on LBW and PTB. It was observed that household income (and other measures of SES) moderates the effect of temperature on birth outcomes and the burden of adverse effects is higher for populations in a socioeconomic disadvantaged situation.

The main objective of this study was to further the understanding of the effects of intrauterine exposure to extreme temperatures on health at birth. Our work contributes to the available literature in various ways. Foremost, it delivers presumably causal estimates of the impact of prenatal exposure to extreme temperatures on a comprehensive set of birth outcomes in the Netherlands. The finding that exposure to extreme high temperatures has a detrimental effect on birth weight and LBW, are in line with results from previous studies.^17–19,21^ We also found that exposure to extreme cold temperatures had a negative impact on LBW and PTB. Although for the latter, the confidence intervals were wide and covered the null, suggesting reduced statistical power, we did however observe a clearer signal for PTB (and LBW) with second-trimester exposure. These results are in line with the conclusions from previous studies.^18,21^ We expanded previous work by investigating whether the observed effect on LBW is related to a preterm delivery (PTB), growth restriction (operationalized as SGA), or both. Given that we observed an effect of exposure to hot days (> 20 °C) for both SGA and PTB, it is sensitive to think that the impact observed on LBW is related to both, preterm deliveries and growth restriction. However, for cold temperatures, an effect was found for PTB (second trimester) but not for SGA. This finding, in addition to the observed pattern that the odds ratios are larger and more consistently deviating from the reference category for PTB points that the effects could be mostly driven by preterm delivery. However, further research into other populations would be required to confirm this statement.

Our study also contributes to the literature by exploring the heterogeneity of the effect of temperature by socioeconomic conditions and fetal sex. Exploring the role of SES as moderator for the effects of exposure to extreme temperatures on birth outcomes can help to provide insights into the potential determinants of disparities in early-life health. It was observed that household income moderated the effect of temperature on birth outcomes and that the detrimental impact of extreme cold and hot days was more sizable for those in disadvantaged socioeconomic circumstances. Differences observed across SES groups could be related to, e.g., physical circumstances of living and working environment, activity patterns, resources available for the adoption of coping strategies or differences in awareness. When looking at the differences by foetal sex, we observed a larger effect for male than for female foetuses. This in line with previous findings that adverse in-utero (environmental) exposures impose larger negative effects on males.^50^ The effect size may seem small at first, representing the impact of a single additional day of exposure. However, considering longer periods, especially for the low SES group, the effects become sizable.

A main strength of this study is its robust approach to investigating the potential effect of exposure to extreme temperatures on birth outcomes. To be able to identify a plausibly causal effect we have leveraged arguably random daily fluctuations in temperature with adjustment for a broad set of fixed effects and climatological variables. Our approach has the advantage over other methods that it can be applied to a wide variety of outcomes regardless of whether they are expected to have an acute-onset or not (as expected in case-crossover analysis). Furthermore, it facilitated the exploration of critical windows of susceptibility. The use of high-quality routinely collected data corresponding to an extended period (2003-2017) led to over 2.4 million individual records being available for analysis. Given its climatological characteristics, the Netherlands provides an ideal research scenario as self-selection into different climate regions is unlikely in this context (which was confirmed by a sensitivity analysis). Another advantage of the study setting is related to the role of adaptation, e.g., individuals in historically hotter places may adapt to high temperatures through the adoption of mitigating technologies, such as air conditioning, or behavioural adaptations.^19^ As mentioned before, extreme temperatures have been rare in the Netherlands during the study period and adaptation strategies to warm weather, such as the use of air conditioning were not widespread throughout the country. In fact, in 2018, only 6% of the Dutch households owned an air conditioner of any sort, and this value can only be lower for the previous years. ^51^ For comparison, in the USA, one of the nations with the highest air conditioning adoption, almost 90% of households in 2015 had air conditioning.^52^ Finally, our results are robust to various specifications as confirmed in the sensitivity analyses.

A limitation of this study is that temperature exposure was based on measurements from the nearest monitor to the residential address of the mother, which may not reflect the temperature in the exact place of residence. This might be particularly relevant for urban populations, who might be exposed to e.g., hotter temperatures than the ones registered in monitor stations due to the urban heat island effect (UHI). The UHI refers to the phenomenon when urban areas experience higher temperature compared to their surrounding non-urban areas,^53^ which has been observed to be more prominent in disadvantaged areas often characterized by a lack of green spaces and poor built environment.^54^ Also, exposure at e.g., the working environment could not be observed along with information on personal activity patterns, such as time spent indoors vs. outdoors. Exposure at the work place and activity patterns might explain at least in part some of the disparities observed across socioeconomic groups. Last, it is likely that our results are an underestimation of the effect of temperature on pregnancy outcomes due to selection, as early exposure to extreme temperatures might lead to spontaneous abortion of fetuses below-average health even before clinical recognition.^17,55,56^ In our analysis we have adjusted our models for common causes of early foetal loss and the outcomes of interest, however, it is likely that this bias cannot be fully addressed. Thus, our estimates should be seen as a lower bound of the true effect.^17,40^

Future research needs to focus on the potential mechanisms through which temperature influences health at birth, particularly those that could be intervened on by public health policy. Previous research has proposed that aside from biological mechanisms, behavioural responses to unusually warm temperatures might also contribute to the effect observed on adverse outcomes.^21,57^ For instance, pregnant women in historically cooler countries might spend more time outdoors when temperatures are unusually warm and engage in more physical activity, potentially raising the risk of fatigue and dehydration.^21,57^ Furthermore, studies aiming at assessing the role of air pollution as a potential mediator (and moderators) for the effect of temperature on health at birth are needed.

Our results are particularly timely and policy relevant, particularly in the light of the recently published State of the Climate in Europe 2022 report,^58^ which highlights Europe as the fastest warming continent in the world. With the frequency of extreme weather events only predicted to increase, public health adaptation strategies for climate change, on a national as well as community level, need to be developed. Furthermore, the identification of vulnerable populations and windows of vulnerability to temperature can assist healthcare providers in constructing and refining the set of recommendations given to pregnant women.

In summary, in this nationwide population-based study in the Netherlands, we found consistent evidence of a detrimental impact of intrauterine exposure to extreme temperatures on adverse birth outcomes, particularly for the exposure during the third trimester. These adverse effects were consistently larger for socioeconomically disadvantaged populations. Thus, the predicted increases in the intensity and frequency of extreme heat episodes has the potential to increase socioeconomic health inequalities at birth.

## Data Availability

This study is based on registry data from the Dutch Perinatal Registry (Perined) and microdata from Statistics Netherlands (https://www.cbs.nl/nl-nl). Access to linked electronic health and sociodemographic records requires approval from Statistics Netherlands following the procedure on their website.

https://github.com/LizBurgosOchoa/Temperature_BirthOutcomes

