## supplementary file for "The impact of extreme temperatures on birth outcomes in the Netherlands: a nationwide population-based study"

Supplementary Files

Supplementary file 1: Full numerical results for the main analysis.

| **Table S1.** Effect of in utero temperature exposure on birth outcomes. [a] | | | |
|---|---|---|---|
| **Temperature bin** | **LBW** [b] | **SGA** [b] | **PTB** [b] |
| < - 4 °C | 1.001 (0.997, 1.004) | 0.999 (0.997, 1.002) | 1.002 (0.999, 1.005) |
| -4 – 0 °C | 0.997 (0.996, 0.999) | 0.998 (0.997, 1.000) | 0.998 (0.996, 1.000) |
| 0 – 4 °C | 0.997 (0.996, 0.999) | 0.999 (0.998, 1.001) | 0.997 (0.996, 0.998) |
| 4 – 8 °C | 0.997 (0.996, 0.998) | 0.999 (0.998, 1.000) | 0.998 (0.997, 0.999) |
| 12 – 16 °C | 1.001 (1.000, 1.002) | 1.001 (1.000, 1.001) | 1.001 (1.000, 1.002) |
| 16 – 20 °C | 1.004 (1.003, 1.005) | 1.001 (1.000, 1.001) | 1.004 (1.003, 1.005) |
| > 20 °C | 1.007 (1.005, 1.009) | 1.004 (1.003, 1.005) | 1.006 (1.005, 1.007) |

LBW, low birthweight; SGA, small for gestational age; PTB, preterm birth.

[a] The effect of in utero exposure to one additional day falling in certain temperature bin on birth outcomes relative to a day with a mean temperature of 8 – 12 °C. [b] For binary outcomes, estimates correspond to Odds Ratios (95% CI) from logistic regression models. All models include province × month fixed effects, province × year-time-trend, and year fixed effects. Environmental controls include mean precipitation, wind speed, sunshine duration, and relative humidity during the gestational period. Other covariates included were maternal age in categories, parity, fetal sex, household income, mother's migration background and education.

Supplementary file 2: Results by gestational trimester.

**Table S2.** Effect of in utero temperature exposure on birth outcomes by gestational trimester, odds ratios (95% CI).[a]

| Temperature bin | LBW [b] | SGA [b] | PTB [b] |
|---|---|---|---|
| **Trimester 1** | | | |
| < - 4 °C | 0.998 (0.993, 1.003) | 0.999 (0.996, 1.002) | 0.997 (0.992, 1.002) |
| -4 – 0 °C | 0.993 (0.991, 0.996) | 0.999 (0.997, 1.000) | 0.995 (0.992, 0.997) |
| 0 – 4 °C | 0.999 (0.997, 1.000) | 0.999 (0.998, 1.000) | 0.999 (0.998, 1.000) |
| 4 – 8 °C | 0.994 (0.992, 0.995) | 0.999 (0.998, 1.000) | 0.994 (0.992, 0.995) |
| 12 – 16 °C | 0.996 (0.994, 0.998) | 1.000 (0.999, 1.001) | 0.996 (0.995, 0.998) |
| 16 – 20 °C | 1.001 (1.000, 1.003) | 1.000 (0.999, 1.001) | 1.001 (0.999, 1.002) |
| > 20 °C | 1.001 (0.999, 1.004) | 1.003 (1.002, 1.004) | 1.001 (0.999, 1.003) |
| **Trimester 2** | | | |
| < - 4 °C | 1.006 (1.001, 1.011) | 0.999 (0.996, 1.003) | 1.003 (1.001, 1.005) |
| -4 – 0 °C | 0.995 (0.993, 0.997) | 0.999 (0.998, 1.001) | 1.000 (0.998, 1.002) |
| 0 – 4 °C | 0.999 (0.998, 1.001) | 1.000 (0.999, 1.001) | 1.000 (0.998, 1.002) |
| 4 – 8 °C | 0.996 (0.995, 0.998) | 0.999 (0.998, 1.000) | 1.001 (0.999, 1.002) |
| 12 – 16 °C | 1.000 (0.999, 1.002) | 1.001 (1.000, 1.002) | 0.999 (0.997, 1.001) |
| 16 – 20 °C | 1.004 (1.002, 1.006) | 1.001 (1.000, 1.002) | 1.003 (1.002, 1.005) |
| > 20 °C | 1.007 (1.006, 1.009) | 1.004 (1.003, 1.006) | 1.003 (1.002, 1.005) |
| **Trimester 3** | | | |
| < - 4 °C | 1.002 (0.997, 1.006) | 1.001 (0.997, 1.004) | 0.992 (0.988, 0.996) |
| -4 – 0 °C | 1.000 (0.998, 1.002) | 0.997 (0.996, 0.999) | 0.999 (0.997, 1.001) |
| 0 – 4 °C | 1.002 (0.999, 1.003) | 1.000 (0.999, 1.001) | 0.998 (0.997, 0.999) |
| 4 – 8 °C | 1.000 (0.998, 1.001) | 0.999 (0.997, 1.000) | 0.997 (0.996, 0.998) |
| 12 – 16 °C | 0.999 (0.997, 1.001) | 1.000 (0.999, 1.001) | 0.997 (0.995, 0.998) |
| 16 – 20 °C | 1.004 (1.002, 1.006) | 1.001 (1.000, 1.002) | 1.001 (1.000, 1.002) |
| > 20 °C | 1.004 (1.003, 1.006) | 1.004 (1.002, 1.005) | 1.005 (1.003, 1.007) |

LBW, low birthweight; SGA, small for gestational age; PTB, preterm birth.
a The effect of in utero exposure to one additional day falling in certain temperature bin on birth outcomes relative to a day with a mean temperature of 8 – 12 °C. b For binary outcomes, estimates correspond to Odds Ratios (95% CI) from logistic regression models. c For continuous outcomes estimates correspond to beta coefficients (95% CI) from linear regression models. All models include province × month fixed effects, province × year-time-trend, and year fixed effects. Environmental controls include mean precipitation, wind speed, sunshine duration, and relative humidity during the gestational period. Other covariates included were maternal age in categories, parity, fetal sex, household income, mother's migration background and education.

Supplementary file 3: Differential effect of temperature exposure across socioeconomic status groups.

**Table S3.** Effect of in utero temperature exposure on birth outcomes by household income groups. [a]

| Temperature bin | LBW [b] | SGA [b] | PTB [b] |
|---|---|---|---|
| **High** | | | |
| < - 4 °C | 0.993 (0.988, 0.997) | 0.989 (0.986, 0.992) | 0.997 (0.993, 1.001) |
| -4 – 0 °C | 0.997 (0.995, 0.998) | 0.998 (0.997, 1.000) | 0.998 (0.996, 0.999) |
| 0 – 4 °C | 1.000 (0.999, 1.001) | 1.000 (0.999, 1.000) | 1.001 (1.000, 1.002) |
| 4 – 8 °C | 0.997 (0.996, 0.998) | 0.999 (0.998, 1.000) | 0.997 (0.996, 0.998) |
| 12 – 16 °C | 1.001 (1.000, 1.002) | 1.000 (1.000, 1.001) | 1.001 (1.000, 1.002) |
| 16 – 20 °C | 1.003 (1.002, 1.005) | 1.000 (1.000, 1.001) | 1.004 (1.003, 1.005) |
| > 20 °C | 1.001 (1.000, 1.003) | 0.998 (0.997, 0.999) | 1.005 (1.004, 1.007) |
| **Medium** | | | |
| < - 4 °C | 0.998 (0.994, 1.002) | 0.997 (0.995, 0.999) | 1.001 (0.998, 1.004) |
| -4 – 0 °C | 0.997 (0.995, 0.998) | 0.998 (0.997, 0.999) | 0.998 (0.996, 0.999) |
| 0 – 4 °C | 1.000 (0.999, 1.001) | 1.000 (0.999, 1.000) | 1.001 (1.000, 1.002) |
| 4 – 8 °C | 0.997 (0.996, 0.998) | 0.999 (0.998, 1.000) | 0.997 (0.996, 0.998) |
| 12 – 16 °C | 1.001 (1.000, 1.002) | 1.000 (0.999, 1.001) | 1.001 (1.000, 1.002) |
| 16 – 20 °C | 1.003 (1.002, 1.005) | 1.000 (0.999, 1.001) | 1.004 (1.003, 1.005) |
| > 20 °C | 1.004 (1.003, 1.006) | 1.002 (1.001, 1.003) | 1.005 (1.004, 1.007) |
| **Low** | | | |
| < - 4 °C | 1.016 (1.011, 1.021) | 1.016 (1.012, 1.019) | 1.009 (1.004, 1.013) |
| -4 – 0 °C | 0.997 (0.995, 0.998) | 0.998 (0.997, 0.999) | 0.998 (0.996, 0.999) |
| 0 – 4 °C | 1.000 (0.999, 1.001) | 1.000 (0.999, 1.001) | 1.001 (1.000, 1.002) |
| 4 – 8 °C | 0.997 (0.996, 0.998) | 0.999 (0.998, 1.000) | 0.997 (0.996, 0.998) |
| 12 – 16 °C | 1.001 (1.000, 1.002) | 1.000 (0.999, 1.001) | 1.001 (1.000, 1.002) |
| 16 – 20 °C | 1.003 (1.002, 1.005) | 1.000 (0.999, 1.001) | 1.004 (1.003, 1.005) |
| > 20 °C | 1.014 (1.012, 1.015) | 1.013 (1.012, 1.014) | 1.010 (1.008, 1.011) |

LBW, low birthweight; SGA, small for gestational age; PTB, preterm birth.

[a] The effect of in utero exposure to one additional day falling in certain temperature bin on birth outcomes relative to a day with a mean temperature of 8 – 12 °C. [b] For binary outcomes, estimates correspond to Odds Ratios (95% CI) from logistic regression models. [c] For continuous outcomes estimates correspond to beta coefficients (95% CI) from linear regression models. All models include province × month fixed effects, province × year-time-trend, and year fixed effects. Environmental controls include mean precipitation, wind speed, sunshine duration, and relative humidity during the gestational period. Other covariates included were maternal age in categories, parity, fetal sex, household income, mother’s migration background and education.

| **Table S4.** Effect of in utero temperature exposure on birth outcomes by neighbourhood socioeconomic status groups. [a] | | | |
|---|---|---|---|
| | **High** | | |
| **Temperature bin** | **LBW** [b] | **SGA** [b] | **PTB** [b] |
| < - 4 °C | 0.994 (0.989, 0.999) | 0.994 (0.991, 0.997) | 1.000 (0.996, 1.005) |
| -4 – 0 °C | 0.997 (0.996, 0.999) | 0.999 (0.998, 1.000) | 0.998 (0.996, 0.999) |
| 0 – 4 °C | 1.000 (0.999, 1.001) | 1.000 (0.999, 1.001) | 1.001 (1.000, 1.002) |
| 4 – 8 °C | 0.997 (0.996, 0.998) | 0.999 (0.998, 1.000) | 0.998 (0.997, 0.998) |
| 12 – 16 °C | 1.001 (1.000, 1.002) | 1.000 (0.999, 1.001) | 1.001 (1.000, 1.002) |
| 16 – 20 °C | 1.003 (1.002, 1.004) | 1.001 (1.000, 1.002) | 1.003 (1.002, 1.004) |
| > 20 °C | 1.001 (1.000, 1.003) | 0.999 (0.998, 1.000) | 1.004 (1.002, 1.005) |
| | **Medium** | | |
| < - 4 °C | 0.997 (0.994, 1.001) | 0.996 (0.994, 0.999) | 1.000 (0.997, 1.004) |
| -4 – 0 °C | 0.997 (0.996, 0.999) | 0.999 (0.998, 1.000) | 0.998 (0.996, 0.999) |
| 0 – 4 °C | 1.000 (0.999, 1.001) | 1.000 (0.999, 1.001) | 1.001 (1.000, 1.002) |
| 4 – 8 °C | 0.997 (0.996, 0.998) | 0.999 (0.998, 1.000) | 0.998 (0.997, 0.998) |
| 12 – 16 °C | 1.001 (1.000, 1.002) | 1.000 (0.999, 1.001) | 1.001 (1.000, 1.002) |
| 16 – 20 °C | 1.003 (1.002, 1.004) | 1.001 (1.000, 1.002) | 1.003 (1.002, 1.004) |
| > 20 °C | 1.004 (1.002, 1.005) | 1.001 (1.000, 1.002) | 1.005 (1.004, 1.006) |
| | **Low** | | |
| < - 4 °C | 1.008 (1.004, 1.012) | 1.009 (1.006, 1.012) | 1.004 (1.000, 1.008) |
| -4 – 0 °C | 0.997 (0.996, 0.999) | 0.999 (0.998, 1.000) | 0.998 (0.996, 0.999) |
| 0 – 4 °C | 1.000 (0.999, 1.001) | 1.000 (0.999, 1.001) | 1.001 (1.000, 1.002) |
| 4 – 8 °C | 0.997 (0.996, 0.998) | 0.999 (0.998, 1.000) | 0.998 (0.997, 0.998) |
| 12 – 16 °C | 1.001 (1.000, 1.002) | 1.000 (0.999, 1.001) | 1.001 (1.000, 1.002) |
| 16 – 20 °C | 1.003 (1.002, 1.004) | 1.001 (1.000, 1.001) | 1.003 (1.002, 1.004) |
| > 20 °C | 1.011 (1.010, 1.013) | 1.008 (1.008, 1.009) | 1.009 (1.007, 1.010) |

LBW, low birthweight; SGA, small for gestational age; PTB, preterm birth.

[a] The effect of in utero exposure to one additional day falling in certain temperature bin on birth outcomes relative to a day with a mean temperature of 8 – 12 °C. [b] For binary outcomes, estimates correspond to Odds Ratios (95% CI) from logistic regression models. All models include province × month fixed effects, province × year-time-trend, and year fixed effects. Environmental controls include mean precipitation, wind speed, sunshine duration, and relative humidity during the gestational period. Other covariates included were maternal age in categories, parity, fetal sex, household income, mother's migration background and education.

| **Table S5.** Effect of in utero temperature exposure on birth outcomes by mother's education groups.[a] | | | | |
|---|---|---|---|---|
| | | **High** | | |
| **Temperature bin** | **LBW** [b] | **SGA** [b] | **PTB** [b] | **Birthweight** [c] |
| < - 4 °C | 1.003 (0.997, 1.008) | 0.997 (0.994, 1.000) | 1.002 (0.998, 1.006) | -0.94 (-1.70, -0.18) |
| -4 – 0 °C | 0.997 (0.995, 0.999) | 0.998 (0.996, 1.000) | 0.997 (0.996, 0.999) | 0.62 (0.44, 0.81) |
| 0 – 4 °C | 1.000 (0.999, 1.001) | 1.000 (0.999, 1.001) | 1.001 (1.000, 1.002) | 0.01 (-0.10, 0.11) |
| 4 – 8 °C | 0.997 (0.996, 0.998) | 0.999 (0.998, 1.000) | 0.997 (0.996, 0.998) | 0.58 (0.45, 0.70) |
| 12 – 16 °C | 1.001 (1.000, 1.002) | 1.000 (0.999, 1.001) | 1.001 (1.000, 1.002) | -0.2 (-0.31, -0.09) |
| 16 – 20 °C | 1.003 (1.002, 1.004) | 1.000 (0.999, 1.001) | 1.004 (1.003, 1.004) | -0.58 (-0.7, -0.47) |
| > 20 °C | 1.006 (1.004, 1.008) | 1.003 (1.002, 1.004) | 1.005 (1.003, 1.007) | -1.08 (-1.35, -0.81) |
| | | **Medium** | | |
| < - 4 °C | 1.000 (0.996, 1.004) | 1.002 (0.999, 1.004) | 1.002 (0.998, 1.005) | -0.97 (-1.43, -0.50) |
| -4 – 0 °C | 0.997 (0.995, 0.999) | 0.998 (0.996, 1.000) | 0.997 (0.996, 0.999) | 0.62 (0.44, 0.81) |
| 0 – 4 °C | 1.000 (0.999, 1.001) | 1.000 (0.999, 1.001) | 1.001 (1.000, 1.002) | 0.01 (-0.10, 0.11) |
| 4 – 8 °C | 0.997 (0.996, 0.998) | 0.999 (0.998, 1.000) | 0.997 (0.996, 0.998) | 0.58 (0.45, 0.70) |
| 12 – 16 °C | 1.001 (1.000, 1.002) | 1.000 (0.999, 1.001) | 1.001 (1.000, 1.002) | -0.20 (-0.31, -0.09) |
| 16 – 20 °C | 1.003 (1.002, 1.004) | 1.000 (0.999, 1.001) | 1.004 (1.003, 1.004) | -0.58 (-0.70, -0.47) |
| > 20 °C | 1.006 (1.005, 1.008) | 1.004 (1.003, 1.004) | 1.005 (1.003, 1.006) | -1.29 (-1.46, -1.12) |
| | | **Low** | | |
| < - 4 °C | 1.000 (0.995, 1.004) | 1.004 (1.000, 1.008) | 1.002 (0.996, 1.007) | -0.39 (-0.88, 0.10) |
| -4 – 0 °C | 0.997 (0.995, 0.999) | 0.998 (0.996, 1.000) | 0.997 (0.996, 0.999) | 0.62 (0.44, 0.81) |
| 0 – 4 °C | 1.000 (0.999, 1.001) | 1.000 (0.999, 1.001) | 1.001 (1.000, 1.002) | 0.01 (-0.1, 0.11) |
| 4 – 8 °C | 0.997 (0.996, 0.998) | 0.999 (0.998, 1.000) | 0.997 (0.996, 0.998) | 0.58 (0.45, 0.7) |
| 12 – 16 °C | 1.001 (1.000, 1.002) | 1.000 (0.999, 1.001) | 1.001 (1.000, 1.002) | -0.20 (-0.31, -0.09) |
| 16 – 20 °C | 1.003 (1.002, 1.004) | 1.000 (0.999, 1.001) | 1.004 (1.003, 1.004) | -0.58 (-0.70, -0.47) |
| > 20 °C | 1.007 (1.005, 1.008) | 1.003 (1.002, 1.005) | 1.007 (1.005, 1.008) | -1.30 (-1.48, -1.12) |

LBW, low birthweight; SGA, small for gestational age; PTB, preterm birth.

[a] The effect of in utero exposure to one additional day falling in certain temperature bin on birth outcomes relative to a day with a mean temperature of 8 – 12 °C. [b] For binary outcomes, estimates correspond to Odds Ratios (95% CI) from logistic regression models. All models include province × month fixed effects, province × year-time-trend, and year fixed effects. Environmental controls include mean precipitation, wind speed, sunshine duration, and relative humidity during the gestational period. Other covariates included were maternal age in categories, parity, fetal sex, household income, mother's migration background and education.

Supplementary file 4: Sensitivity analyses.

**Table S6.** Effect of in utero temperature exposure on birth outcomes additionally adjusted for distance to monitoring station. [a]

| Temperature bin | LBW [b] | SGA [b] | PTB [b] |
|---|---|---|---|
| < - 4 °C | 1.001 (0.997, 1.004) | 0.999 (0.997, 1.002) | 1.002 (0.999, 1.005) |
| -4 – 0 °C | 0.998 (0.996, 0.999) | 0.999 (0.997, 1.001) | 0.998 (0.997, 1.000) |
| 0 – 4 °C | 1.000 (0.998, 1.001) | 1.000 (0.999, 1.001) | 0.999 (0.998, 1.000) |
| 4 – 8 °C | 0.997 (0.996, 0.999) | 0.999 (0.998, 1.000) | 0.998 (0.997, 0.999) |
| 12 – 16 °C | 1.001 (1.000, 1.002) | 1.001 (1.000, 1.001) | 1.001 (1.000, 1.002) |
| 16 – 20 °C | 1.004 (1.003, 1.005) | 1.001 (1.000, 1.001) | 1.004 (1.003, 1.005) |
| > 20 °C | 1.007 (1.005, 1.008) | 1.004 (1.003, 1.005) | 1.006 (1.005, 1.007) |

LBW, low birthweight; SGA, small for gestational age; PTB, preterm birth.
[a] The effect of in utero exposure to one additional day falling in certain temperature bin on birth outcomes relative to a day with a mean temperature of 8 – 12 °C. [b] For binary outcomes, estimates correspond to Odds Ratios (95% CI) from logistic regression models. All models include province × month fixed effects, province × year-time-trend, and year fixed effects. Environmental controls include mean precipitation, wind speed, sunshine duration, and relative humidity during the gestational period. Other covariates included were maternal age in categories, parity, fetal sex, household income, mother's migration background and education.

| **Table S7.** Effect of temperature negative control (placebo) exposure on birth outcomes. [a] | | | |
|---|---|---|---|
| **Temperature bin** | **LBW** [b] | **SGA** [b] | **PTB** [b] |
| < - 4 °C | 0.996 (0.993, 1.000) | 1.000 (0.998, 1.003) | 0.996 (0.993, 0.999) |
| -4 – 0 °C | 0.996 (0.993, 1.000) | 0.999 (0.998, 1.000) | 0.996 (0.994, 0.997) |
| 0 – 4 °C | 0.998 (0.996, 1.001) | 1.000 (0.999, 1.001) | 0.998 (0.997, 0.999) |
| 4 – 8 °C | 0.998 (0.996, 1.001) | 0.999 (0.998, 1.000) | 0.997 (0.996, 0.998) |
| 12 – 16 °C | 0.999 (0.998, 1.000) | 1.000 (0.999, 1.001) | 0.998 (0.997, 0.999) |
| 16 – 20 °C | 0.997 (0.994, 1.000) | 0.999 (0.998, 1.000) | 0.995 (0.994, 0.996) |
| > 20 °C | 1.000 (0.998, 1.001) | 1.000 (0.999, 1.001) | 0.997 (0.994, 1.000) |

LBW, low birthweight; SGA, small for gestational age; PTB, preterm birth.

[a] The effect of placebo exposure (9 months after birth) to one additional day falling in certain temperature bin on birth outcomes relative to a day with a mean temperature of 8 – 12 °C. [b] For binary outcomes, estimates correspond to Odds Ratios (95% CI) from logistic regression models. All models include province × month fixed effects, province × year-time-trend, and year fixed effects. Environmental controls include mean precipitation, wind speed, sunshine duration, and relative humidity during the gestational period. Other covariates included were maternal age in categories, parity, fetal sex, household income, mother's migration background and education.

| **Table S8.** Effect of in utero temperature exposure on birth outcomes using minimum temperature to specify exposure bins. [a] | | | |
|---|---|---|---|
| **Temperature bin** | **LBW** [b] | **SGA** [b] | **PTB** [b] |
| < - 4 °C | 1.001 (1.000, 1.003) | 1.001 (1.000, 1.002) | 1.001 (1.000, 1.002) |
| -4 – 0 °C | 1.000 (0.999, 1.001) | 1.000 (0.999, 1.001) | 1.001 (1.000, 1.002) |
| 0 – 4 °C | 0.998 (0.997, 0.999) | 0.998 (0.996, 1.001) | 0.999 (0.998, 1.000) |
| 4 – 8 °C | 1.001 (1.000, 1.002) | 1.000 (0.998, 1.001) | 0.998 (0.996, 1.000) |
| 12 – 16 °C | 1.002 (1.001, 1.002) | 1.000 (0.999, 1.001) | 1.002 (1.001, 1.003) |
| > 16 °C | 1.003 (1.002, 1.005) | 1.003 (1.002, 1.004) | 1.002 (1.001, 1.003) |

LBW, low birthweight; SGA, small for gestational age; PTB, preterm birth.
[a] The effect of in utero exposure to one additional day falling in certain temperature bin on birth outcomes relative to a day with a minimum temperature of 8 – 12 °C. [b] For binary outcomes, estimates correspond to Odds Ratios (95% CI) from logistic regression models. All models include province × month fixed effects, province × year-time-trend, and year fixed effects. Environmental controls include mean precipitation, wind speed, sunshine duration, and relative humidity during the gestational period. Other covariates included were maternal age in categories, parity, fetal sex, household income, mother's migration background and education.

| **Table S9.** Effect of in utero temperature exposure on birth outcomes using maximum temperature to specify exposure bins. [a] | | | |
|---|---|---|---|
| **Temperature bin** | **LBW** [b] | **SGA** [b] | **PTB** [b] |
| < - 4 °C | 0.998 (0.988, 1.007) | 0.992 (0.986, 0.998) | 1.003 (0.995, 1.012) |
| -4 – 0 °C | 1.003 (1.001, 1.006) | 1.001 (0.999, 1.003) | 1.003 (1.001, 1.006) |
| 0 – 4 °C | 1.000 (0.999, 1.001) | 1.000 (0.999, 1.001) | 1.001 (1.000, 1.002) |
| 4 – 8 °C | 0.998 (0.996, 1.001) | 0.998 (0.996, 1.001) | 0.998 (0.997, 0.999) |
| 12 – 16 °C | 1.000 (0.998, 1.001) | 0.998 (0.996, 1.001) | 1.000 (0.999, 1.001) |
| 16 – 20 °C | 1.000 (0.999, 1.001) | 1.000 (0.998, 1.002) | 1.000 (0.999, 1.001) |
| 20 – 24 °C | 1.001 (1.000, 1.002) | 1.001 (1.000, 1.002) | 1.000 (0.999, 1.001) |
| 24 – 28 °C | 1.007 (1.005, 1.009) | 1.005 (1.004, 1.006) | 1.006 (1.005, 1.008) |
| > 28 °C | 1.009 (1.007, 1.012) | 1.003 (1.002, 1.005) | 1.009 (1.007, 1.011) |

LBW, low birthweight; SGA, small for gestational age; PTB, preterm birth.

[a] The effect of in utero exposure to one additional day falling in certain temperature bin on birth outcomes relative to a day with a maximum temperature of 8 – 12 °C. [b] For binary outcomes, estimates correspond to Odds Ratios (95% CI) from logistic regression models. All models include province × month fixed effects, province × year-time-trend, and year fixed effects. Environmental controls include mean precipitation, wind speed, sunshine duration, and relative humidity during the gestational period. Other covariates included were maternal age in categories, parity, fetal sex, household income, mother’s migration background and education.

**Table S10.** Effect of in utero temperature exposure on birth outcomes by foetal sex. [a]

| Temperature bin | LBW [b] | SGA [b] | PTB [b] |
|---|---|---|---|
| **Male** | | | |
| < - 4 °C | 1.001 (0.996, 1.006) | 1.000 (0.997, 1.004) | 1.003 (0.998, 1.007) |
| -4 – 0 °C | 0.997 (0.994, 0.999) | 0.999 (0.997, 1.001) | 0.999 (0.997, 1.000) |
| 0 – 4 °C | 1.000 (0.999, 1.002) | 0.999 (0.998, 1.000) | 1.002 (1.001, 1.003) |
| 4 – 8 °C | 0.997 (0.995, 0.998) | 0.999 (0.998, 1.000) | 0.999 (0.997, 1.000) |
| 12 – 16 °C | 1.001 (1.000, 1.003) | 1.000 (0.999, 1.001) | 1.002 (1.001, 1.003) |
| 16 – 20 °C | 1.005 (1.004, 1.007) | 1.000 (0.999, 1.001) | 1.006 (1.005, 1.007) |
| > 20 °C | 1.009 (1.007, 1.011) | 1.003 (1.002, 1.005) | 1.007 (1.006, 1.009) |
| **Female** | | | |
| < - 4 °C | 1.001 (0.996, 1.005) | 0.998 (0.995, 1.002) | 1.001 (0.997, 1.006) |
| -4 – 0 °C | 0.998 (0.996, 1.001) | 0.998 (0.996, 1.000) | 0.998 (0.996, 1.000) |
| 0 – 4 °C | 1.000 (0.999, 1.002) | 1.000 (0.999, 1.001) | 1.001 (1.000, 1.002) |
| 4 – 8 °C | 0.998 (0.996, 0.999) | 0.999 (0.998, 1.000) | 0.997 (0.996, 0.999) |
| 12 – 16 °C | 1.000 (0.999, 1.002) | 1.001 (1.000, 1.002) | 0.999 (0.998, 1.001) |
| 16 – 20 °C | 1.002 (1.001, 1.004) | 1.001 (1.000, 1.002) | 1.001 (1.000, 1.003) |
| > 20 °C | 1.005 (1.003, 1.007) | 1.005 (1.004, 1.006) | 1.004 (1.002, 1.006) |

LBW, low birthweight; SGA, small for gestational age; PTB, preterm birth.
[a] The effect of in utero exposure to one additional day falling in certain temperature bin on birth outcomes relative to a day with a mean temperature of 8 – 12 °C. [b] For binary outcomes, estimates correspond to Odds Ratios (95% CI) from logistic regression models. All models include province × month fixed effects, province × year-time-trend, and year fixed effects. Environmental controls include mean precipitation, wind speed, sunshine duration, and relative humidity during the gestational period. Other covariates included were maternal age in categories, parity, fetal sex, household income, mother's migration background and education.
